# Efficacy and effectiveness of hand hygiene-related practices used community settings for removal of organisms from hands: A systematic review

**DOI:** 10.1101/2025.03.12.25323775

**Authors:** Stephen P Hilton, Nick H. An, Lilly A. O’Brien, Jedidiah S. Snyder, Hannah Rogers, Oliver Cumming, Joanna Esteves Mills, Bruce Gordon, Matthew C. Freeman, Bethany A. Caruso, Marlene K Wolfe

## Abstract

**Background:** This systematic review collected and synthesized evidence on the efficacy and effectiveness of commonly used hand hygiene methods for removing or inactivating pathogens on hands in community settings. The evidence was generated to support the development of the WHO Guidelines for Hand Hygiene in Community Settings.

**Methods:** We searched PubMed, Web of Science, EMBASE, CINAHL, Global Health, Cochrane Library, Global Index Medicus, Scopus, PAIS Index, WHO IRIS, UN Digital Library and World Bank eLibrary, and consulted experts. Eligible studies included laboratory and field studies that measured reduction in organisms on hands after washing with hand hygiene products intended for use in community settings; healthcare settings were excluded. Two reviewers independently extracted data from each study; risk of bias was assessed using the Mixed Method Appraisal Tool and a laboratory-based quality assessment tool. Summary results in terms of log reduction in organisms on hands were calculated for categories with five or more data points from two or more studies.

**Results:** Of studies that met inclusion criteria, the majority focused on alcohol-based hand sanitizer (111, 63%) and handwashing with soap and water (110, 62%). Most evidence (66%) assessed bacterial reductions. Commonly used methods showed an overall >2 log reduction in bacteria after handwashing (2.12 [1.91, 2.33] log reduction for soap and water and 3.25 [3.01, 3.49] for alcohol-based sanitizers), but not for viruses (1.57 [1.23, 1.92] for soap and water and 1.72 [1.49, 1.94] for alcohol-based sanitizers).

**Conclusions:** The ability to draw conclusions on efficacy of types of methods was limited methods and organisms studied. Additional evidence quantifying the impact of handwashing in areas with major gaps in available data – data on viruses (especially enveloped viruses), handwashing alternatives other than alcohol-based sanitizers, drying methods and microbial water quality - would support more evidence-based recommendations.

**Funding:** This work was supported by the World Health Organization (PO number: 203046633).

**PROSPERO registration number:** CRD42023429145.

**What is already known on this topic:** Hand hygiene plays a critical role in preventing the spread of infectious disease, however there is a lack of data describing which methods are most efficacious for handwashing in community setting, particularly for handwashing alternatives and handwashing for viruses. Previous reviews have described the effectiveness of handwashing to prevent disease transmission, however, there has not been a thorough review describing the efficacy of handwashing for removing and inactivating organisms.

**What this study adds:** This study provides an overview of the current knowledge on handwashing efficacy and demonstrates that there is substantial data to suggest that both handwashing with soap and water and use of alcohol-based hand sanitizers are efficacious for removing bacteria. However, there are gaps in knowledge around the efficacy of hand hygiene practices for removal and inactivation of viruses and for a wider range of handwashing methods that are commonly practiced around the world such as use of soap alternatives like ash and sand.

**How this study might affect research practice or policy:** Handwashing guidelines across the globe demonstrate inconsistencies and contradictions and lack evidence to support some of the recommended practices. This study provides an overview of the current evidence to support handwashing guidelines and implementation of practices globally and indicates that handwashing with soap and water is typically an efficacious method. However, to formulate strong recommendations for handwashing practices across diverse contexts more data is needed on a wider range of conditions. This review highlights these gaps, including a lack of data on viral pathogens and elements of handwashing practices such as the use of soap alternatives, drying methods, water quality, and timing of washing.

## 1. INTRODUCTION

Hand hygiene, whether through handwashing with soap or other methods such as the use of alcohol-based hand rubs (ABHRs), is a critical behavior to prevent disease transmission. Hand hygiene interventions are relatively inexpensive to implement [1] and can prevent several infectious diseases including enteric and respiratory infections, which account for a large burden of disease [2–4] and high healthcare costs [5]. Establishing global guidelines and recommendations is essential to guide hand hygiene initiatives, protect public health, and strengthen resilient health systems [6].

According to the Ottawa Charter, community settings are where ‘health is created and lived by people within the setting of their everyday life; where they learn, work, play, and love,’ [7] and include domestic, public, and institutional spaces [8]. Guidelines for hand hygiene in healthcare settings are well-established [9–12], and additional guidelines emphasize investing in hand hygiene as a core public health measure [13–16]. Despite the recognized importance of hand hygiene, and the unprecedented prioritization driven by the COVID-19 pandemic [16], gaps and inconsistencies remain in global guidance on specific measures [8]. A recent scoping review identified 51 existing international guidelines and highlighted a lack of consistent evidence-based recommendations and identified four areas where clear recommendations are needed for hand hygiene in community settings: (1) effective hand hygiene; (2) minimum requirements; (3) behavior change; and (4) government measures [8].

Effective hand hygiene refers to hygiene practices that are capable of removing and inactivating organisms from hands. Effectiveness depends on many aspects of the methods used, including the efficacy of handwashing for the removal and inactivation of organisms and the application of these methods in practice in the field. MacLeod et al 2023 found that guidelines for hand hygiene generally agree on what practices are presumed to be effective for hand hygiene; however, it is unclear how evidence-based these guidelines are – i.e. how much data there is to show that these practices are indeed effective. There are many remaining inconsistencies and gaps in the evidence about how to practice hand hygiene to most efficaciously remove and inactivate organisms from hands, and their effectiveness in practice. While guidelines recommend handwashing with soap and water, many hand hygiene methods are available, including the use of products that range from soap to hand sanitizers (both alcohol-based hand rubs and non-alcoholic antiseptics), to antimicrobial wipes and soap substitutes such as ash and sand.

Handwashing with soap and water (or substitutes) is intended to remove organisms from hands, while handwashing with a soap that has added antibacterial properties is intended to remove and inactivate organisms. Use of an antiseptic rub is intended to inactivate organisms but does not wash away organisms or other materials soiling hands. Because the mechanisms of hand hygiene are different among these approaches and may impact different pathogens differently, it is important to establish any differences in efficacy of these hand hygiene options. Among the range of hand hygiene options, the comparative efficacy in removing or deactivating pathogens in community settings is not well-established.

Handwashing efficacy is often investigated using standardized *in vivo* testing methods for evaluating handwashing efficacy in a laboratory, such as standard methods ASTM E2755-15 in the United States and EN1500 in Europe [17,18]. These methods typically involve inoculating a volunteer’s hands with a test organism and then enumerating the organisms on hands before and after handwashing. The results from this testing are usually expressed in terms of log reduction in organisms on hands. Similar testing may also be done in field studies and on naturally contaminated hands, but it is more difficult to enumerate change in such settings. Previous reviews have primarily focused on soap products used as recommended by the WHO [19–21], as well as ABHR [22]. There has been relatively less work done to review efficacious alternatives for hand hygiene when soap or ABHR are not available or are used “imperfectly” [22–25], or for understanding which drying methods prevent recontamination [26].

The aim of this systematic review was to assess the effectiveness and/or efficacy of hand hygiene methods in removing or deactivating pathogens associated with disease transmission in community settings. The priority question and sub-questions for this review were generated through an extensive consultation process by the WHO with external experts [27,28] following a scoping review of current international guidelines [8].

## 2. METHODS

### 2.1. Research questions

This systematic review addressed four questions related to effective/efficacious hand hygiene: (a) How effective are soap products at removing or deactivating key pathogens (or organisms intended as their surrogates) and how does duration impact effectiveness? (b) Where soap and/or water are not available, what are appropriate alternatives for hand hygiene? (c) Which hand-drying methods are effective at reducing risk of recontamination of washed hands? (d) What microbial water quality is required for effective handwashing with soap? In the handwashing literature, laboratory experiments typically provide data on the efficacy of practices and field studies demonstrate their effectiveness in practice – both types of studies were sought in this review as described further below.

### 2.2. Search strategy

This review was pre-registered with PROSPERO (registration number: CRD42023429145) and is reported in accordance with the Preferred Reporting Items for Systematic Reviews and Meta-Analyses (PRISMA) criteria [29] (S1 – PRISMA Checklist). This review was part of an integrated protocol for multiple related reviews to synthesize the evidence for effective hand hygiene in community settings [28]. We adopted a two-phased approach for identifying relevant studies. Phase 1 involved a broad search to capture all studies on hand hygiene in community settings that were relevant across multiple related systematic reviews. The outcome of phase 1 was a reduced sample from which further screening, specific to this review, was performed. A full description of the procedures followed for searches (including search terms), study inclusion, outcomes data collection, analysis, and reporting of the multiple related reviews is presented in the published protocol [28].

This search included studies published between January 1, 1980, and March 29, 2023, and published in English—unless the title and abstract was published in English and/or a non-English language article was referenced in an existing systematic review. We searched 12 peer-reviewed and grey literature databases. PubMed, Web of Science, EMBASE (Elsevier), CINAHL (EBSCOhost), Global Health (CAB), Cochrane Library, Global Index Medicus, Scopus (Elsevier), Public Affairs Information Service (PAIS) Index (ProQuest) were searched on March 23, 2023 and WHO Institutional Repository for Information Sharing (IRIS), UN Digital Library, and World Bank eLibrary were searched on March 28, 2023 using search terms related to hand hygiene broadly and restrictions on terms related to healthcare settings in the titles [28]. We searched trial registries (International Clinical Trials Registry Platform and clinicaltrials.gov) for trials related to hand hygiene in community settings on March 29, 2023.

To ensure an exhaustive search, we conducted manual searches of reference lists of eight relevant systematic reviews [19–26]. For reviews that provided a list of the reviewed articles, we searched only those references. If a list was not available, we searched all references and screened for potentially relevant titles. These reviews had 221 total references of which 120 were duplicates, 33 were already identified in our database search, and 68 were added to phase 2 title and abstract screening. Lastly, we contacted 35 content experts and organizations, using snowballing methods, from April to May 2023 for information on relevant unpublished literature.

### 2.3. Selection criteria

For this review, hand hygiene refers to any hand cleansing undertaken for the purpose of removing or deactivating pathogens from hands and efficacious hand hygiene is defined as any practice which effectively removes or deactivates pathogens from hands and thereby has the potential to limit disease transmission [10]. The term community settings included domestic (e.g., households), public (e.g., markets, public transportation hubs, vulnerable populations [e.g., people experiencing homelessness], parks, squares, or other public outdoor spaces, shops, restaurants, and cafes), and institutional (e.g., workplace, schools and universities, places of worship, prisons and places of detention, nursing homes and long-term care facilities) settings [8]. Studies were excluded if they were in healthcare settings or were animal research. Studies in nursing homes and long-term care facilities were excluded as part of phase 2 screening as these were defined as locations where healthcare is delivered and provided evidence akin to that generated in healthcare settings. There were no geographic restrictions.

Eligible studies were laboratory and field efficacy studies in which hands were either experimentally inoculated or naturally contaminated. Publications not based on empirical research were excluded. Eligible interventions were handwashing with soap and water at varying durations; other handwashing materials including antiseptics, friction-generating materials, and water alone for varying durations; any hand drying method after handwashing with water or soap and water; and hand washing with soap and water contaminated with microorganisms (naturally or experimentally). To meet the eligibility criteria, interventions needed to be in general population in community settings or laboratory-based studies on interventions used in community settings (i.e. surgical scrubs and similar were excluded). Data was also excluded if it represented results after multiple rounds of handwashing, including intermediary steps from handwashing to sample recovery that might affect microbial outcome, or used gloved hands or focused on fingernails only. Studies included reported results of handwashing and drying using the measurement of an appropriate organism before and after handwashing and expressed in terms of log or percent reduction/increase in organisms on hands.

Results were stratified by hand hygiene method, type of organism, and elements of handwashing practice such as time spent washing. For hand hygiene method, handwashing with soap and water included any soap-based product (with or without antimicrobial additives) that was used in combination with water to wash hands, alcohol based rubs included any alcohol-based product that did not require water, non-alcoholic antiseptic included any antiseptic product with a non-alcohol based primary ingredient that did not involve water, and soap alternatives included any non-antiseptic and non-alcoholic product or material that was used for washing with water. Results were also considered based on the type of organism used for testing: bacteria (gram-positive or gram-negative), virus (enveloped or non-enveloped), and fungi.

We used Covidence software for systematic reviews [30]. In both phases, screening of each article (phase 1 – title and abstract only; phase 2 – title and abstract, then full text review) was performed independently by two reviewers, with discordance between reviewers reconciled by a third reviewer. The stages of the search and screening process are described in the PRISMA flow diagram (S2 – PRISMA Flow Diagram).

### 2.4. Data analysis

Two reviewers [KF, NA] independently extracted data using customized data extraction tools (S3 – Covidence Extraction Template and S4 – Outcome Extraction Template) and assessed risk of bias for each article using the Mixed Method Appraisal Tool (MMAT) [31,32]. Relevant studies were also assessed using a tool to evaluate the quality of laboratory-based microbiologic studies developed by Yeargin, et al [33]. Any conflicts between reviewers over data extraction and bias assessment were resolved by discussion and agreement.

We extracted summary-level results (i.e., log or percent reduction/increase) of handwashing and drying using the measurement of an appropriate organism before and after handwashing and/or drying. Results were categorized by handwashing and/or drying practices (e.g., handwashing with soap, use of antiseptics, material for drying) by the type of microorganism studied (bacteria, enveloped viruses, non-enveloped viruses), duration of handwashing, and whether rubbing of hands was part of the handwashing method. Results were synthesized for categories with five or more data points from two or more studies by performing a meta-analysis of studies that reported a log or percent reduction within categories of handwashing practice along with a measure of spread. Studies were also required to specify the type of organism tested and handwashing methods used. Studies that reported concentrations before and after handwashing and did not compute a reduction value, those with results shown just in a figure, and studies that did not provide useable estimates of uncertainty were unable to be included in meta-analysis. For studies with missing log reduction or spread data, we contacted the corresponding author to request that the missing items be shared for inclusion.

Analysis was conducted in R (version 4.3.1), using the “meta” package. Prior to meta-analysis, results expressed in terms of percent reduction were transformed to log reduction values using a Wilson confidence interval approach to estimate the confidence interval. In cases where the upper limit of the confidence interval was 100% pathogen removal, a value of 5 log_10_ reduction was imputed. To generate summary log reduction values, the inverse-variance method was used (which was the default for random models in that R package), with the restricted maximum-likelihood method being used to estimate tau^2^, and with Hartung-Knapp adjustments for random effects models. The variability in outcomes is described by the I^2^ value; the higher the value, the more of the variation is described by heterogeneity among the included results. The quality of evidence for an association between hand hygiene methods and their efficacy at removing or deactivating pathogens were evaluated in accordance with the Grading of Recommendations Assessment, Development and Evaluation system [34].

### 2.5. Ethics and Patient/ Public Involvement Statement

Because we reviewed published documents, no ethical approval was required. Patients or the public were not involved directly in the design, or conduct, or reporting, or dissemination plans of our research. This evidence synthesis supports the forthcoming WHO Guidelines for Hand Hygiene in Community Settings; the study questions were developed in broad consultation with a network of key partners. Findings from this review will be disseminated alongside the Guidelines.

## 3. RESULTS

### 3.1. Characteristics of the studies included in this review

We identified 177 manuscripts that met inclusion criteria. Of these, 160 (90%) reported on laboratory-based studies and 18 (10%) reported on field-based studies, including one manuscript that reported both study types (Table 1). A large percentage of studies (62%, n=110) included data on handwashing with soap and water. A similar number of studies included data on alcohol-based hand rubs (63%, n=111). Fewer manuscripts reported studies on handwashing with water only (18%, n=32), non-alcoholic antiseptics (14%, n=25), handwashing with soap alternatives (8%, n=15), hand drying (7%, n=12), and handwashing with antimicrobial wipes (6%, n=10). An overview of all included manuscripts and their characteristics including MMAT assessment results and laboratory quality assessment results are listed in S5 – Included Studies.

**Table 1.** Characteristics of the included studies.

| Descriptive characteristics of studies | Total<br>n (%) |
| --- | --- |
| **Total number of studies** | **177 (100%)** |
| **Study Type\*** |  |
| General population in community studies | 18 (10%) |
| Laboratory-based studies on interventions used in community setting | 160 (90%) |
| **Hand hygiene practice\*** |  |
| Handwashing with soap and water | 110 (62%) |
| Handwashing with water only | 32 (18%) |
| Handwashing with duration | 100 (59%) |
| Handwashing with microbial water quality | 2 (1%) |
| Hand rubbing with alcohol-based hand rub | 111 (63%) |
| Hand rubbing with non-alcoholic antiseptics | 25 (14%) |
| Handwashing with soap alternatives | 15 (8%) |
| Hand rubbing with antimicrobial wipes | 10 (6%) |
| Hand drying | 12 (7%) |
| **Study quality rating (mean)** |  |
| MMAT | 4.2 |
| Laboratory Quality Assessment | 4.81 |

*\*Descriptive statistics for study setting and hand hygiene practice represent variables where multiple options could be applicable, so the sum of sections may be over the study total.*

### 3.2. Quality of the studies included in this review

Overall, the mean study quality was 4.2 by the MMAT Assessment, and 4.81 by the laboratory assessment (n = 177). Full MMAT quality appraisal scores and laboratory quality scores for each included study are available in S6 – Bias Assessments.

### 3.3 Handwashing with soap and water

A total of 110 (62%) studies reported results related to the effectiveness of soap products for removing and deactivating key pathogens (Table 1). From these, 32 papers yielded 119 data points on bacteria, four papers yielded 25 data points on viruses in the laboratory, and two papers yielded 25 data points on bacteria from the field (Table 2, S7 – Expanded Tables). From laboratory experiments with bacteria there were 43 data points on plain soap, 41 data points on soap with antimicrobial ingredients, and 10 data points on water alone. Data was insufficient to summarize by pathogen type or soap type for viruses or for field studies.

**Table 2.**
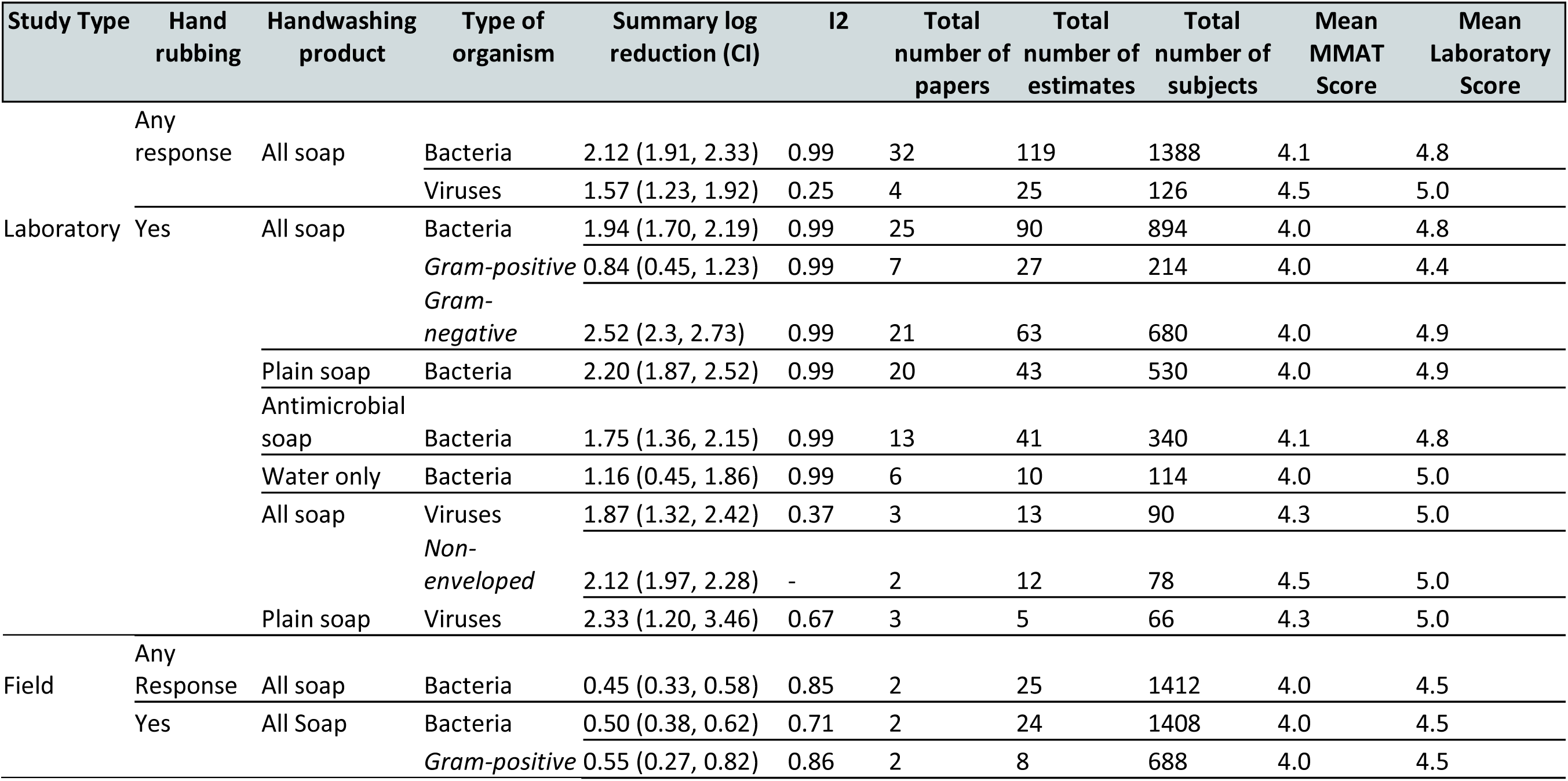
Meta-analysis of studies reporting results on handwashing with soap and water. This table only shows comparisons where data were available.

Washing with soap and water resulted in a summary log reduction value (LRV) for bacteria of 2.12 (95% CI 1.91, 2.33); summary LRVs for bacteria ranged from a low of a 0.84 (95% CI 0.45, 1.23) for gram-positive bacteria to a high of a 2.52 (95% CI 2.30, 2.73) for gram-negative bacteria. For bacteria there did not appear to be a difference in LRV depending on whether the soap used was antibacterial (1.75, 95% CI 1.36, 2.15) or not (2.20, 95% CI 1.87, 2.52) based on substantial overlap in the confidence intervals associated with these estimates (Table 2). For viruses, data revealed a 1.57 (95% CI 1.23, 1.92) summary LRV for handwashing with soap and water; however, when analysis was restricted to studies using plain soap and water for viruses, we found a summary LRV of 2.33 (95% CI 1.2, 3.46). For handwashing with water only, there were six studies with data that could be summarized for bacteria, revealing a summary LRV of 1.16 (95% CI 0.45, 1.86). There was insufficient data for meta-analysis on handwashing experiments that did not involve rubbing hands. For field studies, LRVs ranged from 0.45 (95% CI 0.33, 0.58) for any soap and water methods for bacteria to 0.55 (95% CI 0.27, 0.82) for soap and water with rubbing for gram-positive bacteria (Table 2). Results from the field were insufficient to summarize for viruses. Heterogeneity was high among all groups, with I^2^ ranging from 0.67 - 0.99 with the exception of any soap and water conditions for viruses, which had an I^2^ of 0.25, or 0.37 for when studies explicitly stated that hands were rubbed.

#### 3.3.1 Duration of Handwashing with Soap and Water

A total of 100 studies reported information on the duration of handwashing with soap and water (Table 1). From these, there was enough data for meta-analysis related to bacteria over time periods of 10-20 seconds (21 data points from 6 studies) and 30+ seconds of washing (60 data points from 20 studies) (Table 3, S7 – Expanded Tables). The largest number of studies reported on handwashing for 30+ seconds. Summary LRVs ranged from a minimum of 1.89 (95% CI 0.97, 2.81) for handwashing with antibacterial soap for 10-20 seconds to 2.40 (95% CI 1.87, 2.94) for handwashing for 10-20 seconds with plain soap. However, there was very little difference among log reduction estimates of bacteria across the time categories; the lowest and highest summary LRV estimates have substantial overlap in their confidence intervals. Heterogeneity was also high, with I^2^ ranging from 0.95 - 0.99.

**Table 3.** Meta-analysis of studies reporting results on the duration of handwashing for removal of bacteria with soap and water.

| Study Type | Handwashing Duration | Handwashing product | Type of organism | Summary reduction (CI) | logI2 | Total number of papers | Total number of estimates | Total number of subjects | Mean of MMAT Score | Mean Laboratory Score |
| --- | --- | --- | --- | --- | --- | --- | --- | --- | --- | --- |
| Laboratory | 10-20 seconds | Any soap | Bacteria | 1.91 (1.43, 2.38) | 0.99 | 6 | 21 | 184 | 4.5 | 4.8 |
|  |  | Plain soap |  | 2.4 (1.87, 2.94) | 0.95 | 5 | 7 | 118 | 4.4 | 5.0 |
|  |  | Antibacterial soap |  | 1.89 (0.97, 2.81) | 0.98 | 3 | 8 | 42 | 4.3 | 4.7 |
|  |  | Water only |  | 0.64 (-0.38, 1.65) | 0.98 | 3 | 6 | 63 | 4 | 5 |
|  | 30 seconds + | Any soap | Bacteria | 1.97 (1.65, 2.30) | 0.99 | 20 | 60 | 637 | 4.0 | 4.8 |
|  |  | Plain soap |  | 2.32 (1.92, 2.72) | 0.99 | 16 | 32 | 376 | 3.9 | 4.9 |
|  |  | Antibacterial soap |  | 1.56 (1.06, 2.07) | 0.99 | 10 | 28 | 261 | 4.0 | 4.8 |

### 3.4 Handwashing with antiseptics and soap alternatives

Studies reporting the effectiveness of alternatives to soap and water included any methods for removal or inactivation of products assessed alcohol-based antiseptics (111 studies), non-alcoholic antiseptics such as chlorhexidine, chlorine, and iodine (25 studies), soap alternatives (15 studies), and antimicrobial wipes (10 studies) for removing and deactivating key pathogens (Table 1). Studies eligible for meta-analysis yielded 117 data points on bacteria and 93 for viruses (33 with rubbing, and 51 without) using alcohol-based antiseptics, 52 data points on bacteria and 12 on viruses using non-alcoholic antiseptics, 11 on bacteria using soap alternatives, and 12 data points on bacteria and 1 on viruses using antimicrobial towels. The most data were available for alcohol-based antiseptics. Much of the data on viruses was focused on non-enveloped viruses; we were not able to generate any summary LRVs for enveloped viruses only. Results of meta-analysis ranged from 1.31 (95% CI 0.91, 1.7) summary LRV for non-enveloped viruses with alcohol-based antiseptics to 3.34 (95% CI 3.07, 3.61) summary LRV for gram-negative bacteria with alcohol-based antiseptics (Table 4, S7 – Expanded Tables). Overall, estimates of log reduction were imprecise and did not indicate clear differences in log reduction among groups, with the exception of the difference between bacteria and viruses for alcohol-based antiseptics (Table 4).

**Table 4.**
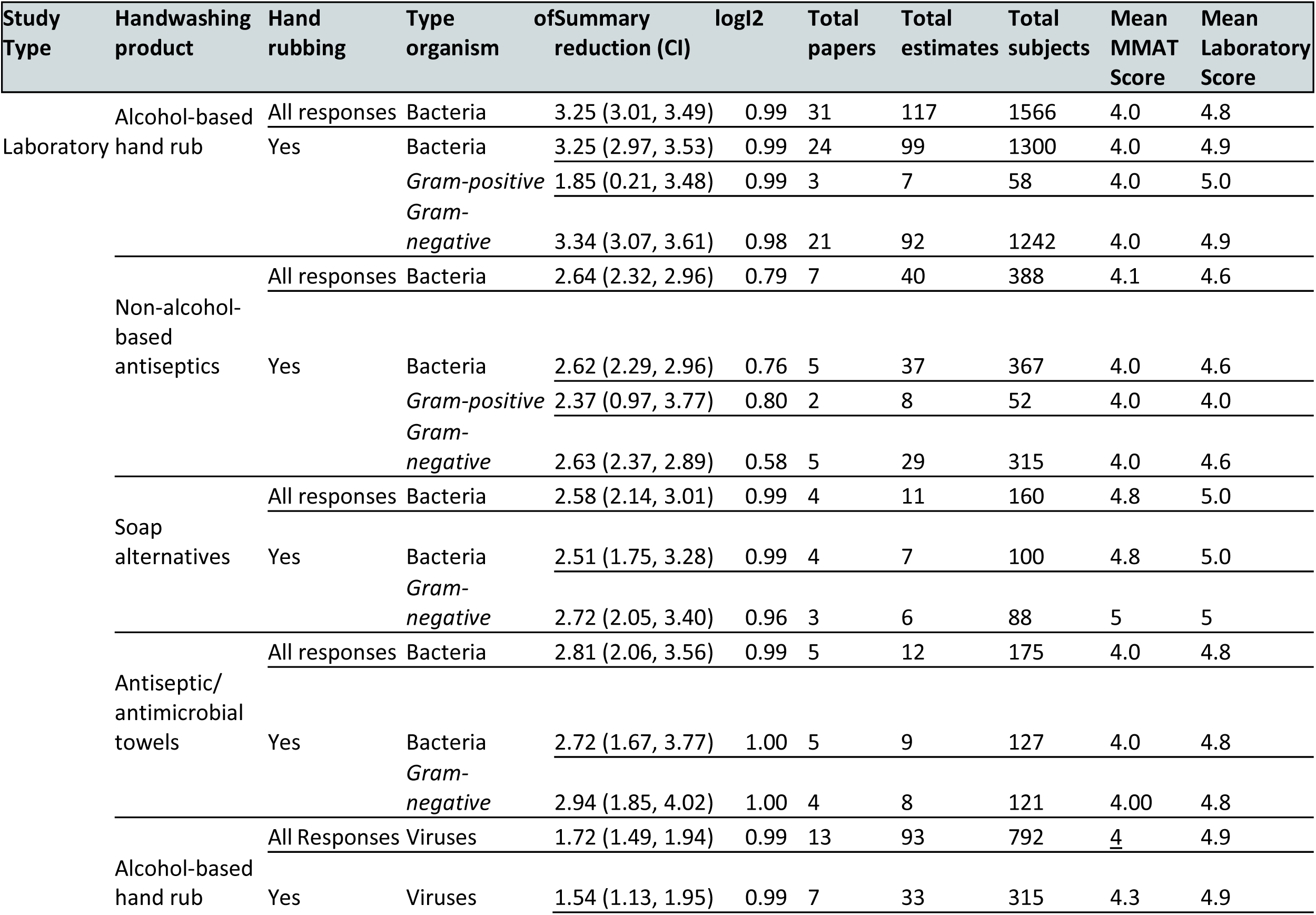
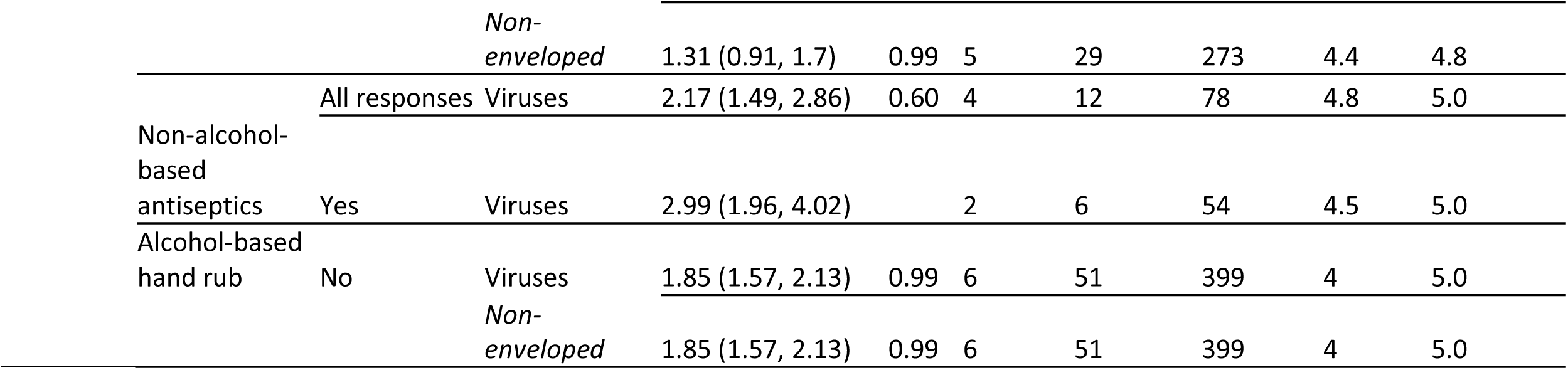
Meta-analysis of studies reporting results on handwashing with soap alternatives.

While soap alternatives as a whole resulted in a 2.58 summary LRV (95% CI 2.14, 3.01), data eligible for meta-analysis was limited to four studies and reported practices differed widely (Table 4). Laboratory studies on soap alternatives reported on the use of ozonized/ozonated water (4 studies), sand (3 studies) ash (3 studies) and *Moringa oleifera* (2 studies) and similar products (S8 – Summary of estimates). Studies on ozonized/ozontaed water found substantial reduction in bacteria on hands, however some found a higher reduction from traditional soap or antiseptics (Breidablik 2020, Appelgrein 2016), while others did not find a significant difference (Nakamura 2021, Breidablik 2019). For sand, Schürmann 1985 found that use of sand resulted in a higher reduction of enteroviruses compared to soap and water (but no statistical comparison was applied), and Zambrana 2023 found that both sand and ash resulted in a LRV similar to soap and water and water only when all were used for a full 20 sec. Both studies on *M. oleifera* use found it ineffective for handwashing in multiple preparations, though one (Torondel 2014) found that a larger volume of powder did result in reductions similar to soap and water (Clark 2018, Torondel 2014).

A total of 10 studies on antimicrobial towels included both single use and reuseable towels with results primarily on bacteria (9 studies) with two studies including viruses and one with fungi (S5 – Included Studies). For bacteria, data summarized from 5 studies demonstrated a summary LRV of 2.81 (2.06, 3.56). Heterogeneity was high (I^2^ =0.99) (Table 4).

### 3.5 Hand drying methods

Overall, 12 studies reported on the impact of drying methods on hand contamination, however most lacked the data necessary for inclusion in a meta-analysis (S5 – Included Studies). Included studies investigated hand drying using paper towels, cloth towels, evaporation, hot air dryers, and jet air dryers. These studies vary in which drying methods are identified as resulting in the greatest decrease or lowest rate of recontamination of hands. While studies that tested paper towels generally reported reductions in contamination (S9 – Drying meta-analysis), no method was consistently identified as showing the greatest reduction across studies. Suen 2019 was the only paper to include all the information required for inclusion in meta-analysis, however this was not carried out because they are from a single study. They performed a laboratory experiment on the effects of six hand drying methods and found that jet air dryers were the most effective in reducing microbial loads on washed hands.

### 3.6 Microbial water quality

Two studies assessed microbial quality of water for handwashing, and neither investigated the use of soap (S10 – Water quality meta-analysis). Yoko et al 2014 investigated the impact of water of poor microbial quality on handwashing in a field study in Nepal; they found that handwashing was performed without soap while hands were highly contaminated and concluded that handwashing with poor quality water did not impact bacterial removal. Torondel et al 2021 conducted a laboratory experiment to determine efficacy of the Supertowel (a reusable microfiber towel with antimicrobial properties) and found that there was no difference in efficacy when it was used with water contaminated with *E. coli*.

## 4. DISCUSSION

This systematic review aimed to assess the efficacy and effectiveness of a wide range of handwashing methods used in community settings globally. We considered any material used for handwashing (e.g. soap and water, alcohol-based sanitizer, or non-traditional materials), and assessed differences in the removal of organisms from hands. We found that common methods used and studied for handwashing typically meet standards for efficacious hand hygiene. However, despite the substantial body of evidence on the efficacy of handwashing with soap and water for removal of bacteria, there is a lack of data on handwashing methods that don’t meet WHO recommendations for handwashing (i.e. reduced duration of handwashing, use of alternatives such as ash and sand or water only), and how much less efficacious these methods might be. The majority of studies focused on bacteria, followed by a significant number of studies on viruses – but there is little to no data on other pathogens such as fungi and protozoa. The majority of studies included in the review were studies of laboratory efficacy; data from field studies describing effectiveness for removal and inactivation of organisms was limited, and laboratory studies investigating methods less commonly used or imperfectly used were also rare.

We found that the most commonly recommended handwashing methods – handwashing with soap and rubbing alcohol-based antiseptics - resulted in a >2 log reduction in organisms on hands after handwashing in laboratory-based experiments for bacteria. Although there are no standard criteria for the efficacy of handwashing in community settings [35], we adopted for comparison a threshold of 2 log describing efficacious handwashing as has been applied in both clinical settings [36] and food safety settings [37] by the United States Food and Drug Administration, since these are both settings with greater regulations. Standard test methods for evaluating antiseptics often look at a comparison of the performance of a reference antiseptic wash to establish efficacy, however, these are challenging to compare across studies and don’t apply to studies on soap or soap alternatives. Thus, using a 2-LRV threshold we found that the majority of methods commonly used for handwashing meet standards for efficacious hand hygiene. However, study results were highly heterogeneous and the data on handwashing with soap and water for viruses was especially limited. There was generally more data available for bacteria than for viruses, and there is a particular lack of data for enveloped viruses. This is of concern as enveloped viruses, such as SARS-CoV-2, have caused significant outbreaks in recent years, and there are potential differences in the effectiveness of hand hygiene for different classes of organisms.

### Handwashing with soap

Handwashing with soap and water resulted in a >2 summary LRV for experiments focused on bacteria, and in some cases for viruses. While there did appear to be a difference in log reduction with handwashing with soap and water between bacteria or viruses (2.12 vs 1.57, respectively), this was no longer the case when the analysis was restricted to plain soap. This data for viruses was limited to only four studies and was highly heterogenous. This limited data suggesting viruses may be removed and inactivated by handwashing less efficaciously than bacteria means it is especially important that further research is done on viruses to determine efficacy across practices more robustly. Few studies investigated the efficacy of handwashing with water alone, and data could only be summarized for bacteria. It is critical to generate more data on viruses and handwashing with and without soap to provide better information on what aspects of these practices most impact efficacy of handwashing.

Meta-analyses found plain soap to be similarly effective to antimicrobial soap for removal and inactivation of bacteria, with a slightly higher estimated LRV for plain soap. Summary LRVs were similar across both plain (2.20, CI = 1.87, 2.52) and antimicrobial (1.75 CI = 1.36, 2.15) soap, and the lack of advantage from antimicrobial soap is not surprising. Previous studies have failed to show any advantage conferred by antibacterial soap compared to plain soap; as such many antibacterial additives have subsequently been banned in the United States [38,39]. Summary LRVS were substantially lower, however, for gram-positive bacteria with the use of any soap (0.84, CI = 0.45, 1.23). This emphasizes the importance of considering the type of pathogen in efficacy studies. Results comparing duration of handwashing for bacteria were inconclusive; there was no substantial difference between the two different time categories with data, and there was no summary data available for the middle category, 20-30 seconds of handwashing.

### Soap alternatives

For laboratory based studies, the results of our meta-analysis showed that hand-rubbing with alcohol-based sanitizers resulted in a very high summary LRV (3.25, 95% CI 3.01, 3.49) across all studies of bacteria, but a much lower summary LRV for viruses (1.72, 95% CI 1.49, 1.94) and an even lower value when only non-enveloped viruses were examined (1.31, 95% CI 0.91, 1.70). Alcohol-based hand sanitizer is the most commonly recommended alternative to soap and water for handwashing, but other options exist, including non-alcohol-based antiseptics, soap alternatives like ash and sand, and antimicrobial towels and wipes. Depending on the active ingredients and application, these products may be more or less effective at inactivating different organisms. For example, non-enveloped viruses (such as norovirus) have been shown to be more difficult to inactivate using alcohol-based sanitizers. More work is needed to investigate the efficacy of alcohol-based sanitizers against enveloped viruses, which are expected to be more susceptible to inactivation but did not have enough data in this review for us to generate a summary LRV. Summary results for non-alcohol-based antiseptics resulted in a >2 LRV for both bacteria and viruses suggesting that a number of antiseptic formulations are efficacious.

Data on soap alternatives and wipes also resulted in a >2 summary LRV for bacteria, but with few data points and highly heterogeneous practices, these summary results should be treated with skepticism. More research investigating the effectiveness of alternatives to soap and water on a range of organisms is needed to make confident recommendations. Soap alternatives studied included practices around which there are discordant recommendations among handwashing guidelines such as the use of ash and sand, which are recommended for use in 14% of handwashing guidelines but advised against in 6% of guidelines globally. Results from the papers included in this study showed that ash and sand were both efficacious for handwashing in some conditions, however there were only two studies identified each for ash and sand. While they may not leave hands looking or feeling cleaner, more research on sand and ash may be particularly useful to inform whether these are effective against organisms in resource constrained areas.

### Hand drying

There was little consistency in results for hand drying, and most studies lacked the information required to generate a summary log reduction value describing the impact of different drying methods. Overall, studies tended to report that paper towels were efficacious for reducing contamination and preventing recontamination, which is consistent with findings from a previous systematic review of hand drying methods (Huang et al 2012). However, both that review and this one found that there is no clear advantage to paper towels over other hand drying methods, and that there was substantial disagreement among studies. Part of this may be a result of different levels of moisture remaining on hands during sampling, as wet hands are more likely to retain bacteria.

### Field based studies

Field based studies showed substantially lower log reduction values than laboratory studies, which is expected when hands are naturally contaminated. It is challenging to produce studies on handwashing effectiveness in the field because a high level of contamination beyond what is normally present is needed to quantify organisms on hands both before and after washing, especially if researchers are hoping to demonstrate a > 2 log reduction in these organisms. These data are also difficult to draw direct comparisons from, as there are many factors that vary when handwashing in a real-world setting. Nonetheless, more handwashing data from field settings is needed to demonstrate which factors most impact effectiveness when handwashing is practiced in context.

### Strengths and Limitations

This review was part of an integrated protocol for multiple related reviews which included an exhaustive search strategy encompassing multiple databases and grey literature sources and a two-phased approach to identify relevant literature of hand hygiene in community settings. Yet despite this rigorous approach and the substantial amount of data included in this review, there are also some key limitations. Data in this review is primarily from laboratory-based studies that approximate real world conditions but may not perform the same in reality, limiting conclusions we could make related to filed studies. More work to assess the likely effectiveness of these methods when used in real-world situations can help provide more evidence for recommendations.

The results are highly heterogeneous within all categories, and conclusions for categories based on relatively few studies and data points should be considered carefully. More data is needed to make comparisons of efficacy across different handwashing methods, given the heterogeneity within each category. This is especially true for some of the less commonly studied types of pathogens, such as enveloped viruses, and less commonly studied handwashing methods, such as soap alternatives. Data on handwashing with water only, handwashing for shorter than recommended durations, and with water of poor microbial quality was limited.

## 5. CONCLUSION

This systematic review describes current evidence on handwashing efficacy for the removal and inactivation of organisms in community settings, and it demonstrates that while recommended handwashing practices with water and soap are well described for the removal of bacteria, there are significant gaps in the data on effectiveness of alternative practices and handwashing conditions that are common in real-world community settings. In general, handwashing with soap and water and alcohol-based sanitizers are effective against bacteria, but the data for effectiveness against viruses is limited and less certain in demonstrating effectiveness. To formulate strong recommendations for handwashing practices, particularly considering viral pandemic illnesses and community resource restrictions, further research that describes the effectiveness of a wider range of practices is critical.

## Authors’ contributions

OC, JEM, and BG conceived the review and designed the specific research questions. BAC and MW are the guarantors of the review. MKW conceived and designed the specific analysis strategy described here. LAO and NA led design of the data extraction process with MKW and JSS support. HR led the literature search, and LAO and NA supported the screening process. NA conducted data extraction with support from SH. SH led the analysis, supported manuscript writing, and oversaw revisions. MW led manuscript writing, and NA and SH drafted portions of the manuscript. All authors read and approved the final version of the manuscript.

## Data Availability

All data and extraction templates will be made publicly available upon publication at Figshare (https://doi.org/10.6084/m9.figshare.28370069).

## Acknowledgements

The authors would like to extend their gratitude to all individuals and institutions whose contributions made this systematic review possible. Special thanks to the team of screeners who contributed substantially to the review process as part of phase 1 and phase 2 screening (Rina Das, Shahreen Hussain, Rosemary Madaki, Jordan Honeycutt, Erika Canda, Erin LaFon, Norah McKinley, Josef Zhao, Kainalu Bailey, Michael Horner-Ibler). We are also grateful to the authors of the studies included in this review for their important contributions to the field.

## Patient Involvement Statement

Patients or the public were not involved directly in the design, or conduct, or reporting, or dissemination plans of our research. This evidence synthesis supports the forthcoming WHO Guidelines for Hand Hygiene in Community Settings; the study questions were developed in broad consultation with a network of key partners. Findings from this review will be disseminated alongside the Guidelines.

